# The role of Construction of Healthcare Consortium on the allocation of human resources for primary care resources and its equity in China

**DOI:** 10.1101/2024.05.23.24307796

**Authors:** Shijie Li, Changze Liao, Shengli Zhang

## Abstract

**Objectives:** This study aims to measure the effect of Construction of Healthcare Consortium (CHC) on the allocation and equity of human resources (HR) for primary health care (PHC) in China, at the same time, it provides some data to support the government’s policies improvement in the next stage.

**Methods:** Changes in the equity of allocation of HR for PHC by population are demonstrated through a three-stage approach to inequality analysis that includes the Gini coefficient (G), the Theil index (T), the concentration index (CI) and concentration curve. The GM(1,1) model is used to project the trend of resources from 2021 to 2030.

**Results:** Volume of HR for PHC accelerates at an average rate of growth following the release of CHC in the 2016. whilst some regions have seen their G and T rise between 2012 and 2016, their levels of inequality of allocation for resource shave gradually declined in the years following 2016, but there are exceptions, with the regions of northeast and northwest seeing the opposite. Eastern and northern region account for a larger contribution to intra-regional inequality. Concentration indices and concentration curves indicate that HR for PHC is related to economic income levels. GM (1, 1) projects an increasing trend in resources from 2021 to 2030, but with variations in average growth rates in different regions.

**Conclusions:** The inequality of human resources for primary health care in China is low, however, the inequality between regions has not been eliminated. We still need to take a long-term view to monitor the impact of CHC on the allocation of HR for PHC and its equity in China.

## 1. Introduction

Equity in population health is an important component of social equity [**Error! Reference source not found**.], yet it cannot be ignored that inequities in population health are still widespread around the world, not only in most developing countries, but also in many developed countries [2-4]. If we want to solve the inequality in population health, how to realize the rational distribution of health resources in different regions and different income groups from a multidimensional perspective will become particularly important, because it is one of the important links to improve the inequality in population health [5].

It can be argued that primary health care (PHC) system is the cornerstone of the effective functioning of health systems worldwide [6] and is recognized as the most cost-effective strategy for improving the health of populations[7,8]. After the market-oriented reform of Chinese health care system at the end of the last century, the potential of PHC’s system in China was suppressed for a long period of time, and instead tertiary comprehensive health care institutions overcrowded with patients and health care resources [9]. The operational efficiency of Chinese health care system was compromised by this irrational structure of supply and demand for health services, which also exacerbated population health inequalities to some extent [10]. In 2009, State Council of China launched a new round of healthcare reform to improve the irrationality of Chinese health care system [11]. In 2015, Hierarchical Diagnosis and Treatment System (HDTS) adapted to China’s national conditions was explicitly proposed and regarded as an effective way to realize the new healthcare reform [12]. HDTS strongly emphasizes the place of PHC in improving health equity for populations. Therefore, in order to better implement the strategy of HDTS, as well as to activate the vitality of PHC more vigorously, China National Health Commission put forward a policy called Guiding Opinions on Carrying Out Pilot Construction of Healthcare Consortia (CHC) in 2016 [13]. One of the objectives of this policy is to improve the scarcity and unequal allocation of PHC’s resources in the past, by directing more and better health care resources, especially human resources (HR) for PHC. Taking 2016 as a starting point, this strategy has been implemented for some time, and it is now time to assess their effectiveness. We want to know if it has had a positive impact on the resourcing and equity of PHC, as it sets out to do.

Reviewing the previous studies, Gini coefficient (G), Concentration index (CI) and Concentration curves, Thiel index (T), index of degree of health resource agglomeration (HRAD), etc. were employed in many studies in evaluating the configuration and inequality of health resources [**Error! Reference source not found**.,14,15]. In addition to assessing equity, Zhang also assessed the efficiency of resource allocation in PHC from an input-output perspective [16]. Some scholars have also analyzed the extent to which the distribution of health resources was spatially equitable in terms of geographic accessibility [17]. In addition, there are studies that show possible future trends in health resources through predictive modeling, the results of which may often be able to guide the work of the government to some extent [18,19].

Based on a review and summary of the literature, we chosen the five years before and after the release of policy of CHC as the time period for our study, focused on changes in the allocation of HR for PHC by population and its equity in China. Changes are demonstrated through a three-stage approach to inequality analysis. In the first phase, G was used to measure inequality over time in the allocation of HR for PHC by population; the second stage was to compute T to explore the root of inequality; CI was employed to understand whether resources tend to be allocated to areas with lower or higher incomes, in third stage. It is noteworthy that scientific predictive values are of real value for policy improvement, yet relatively few studies have been conducted in the past to predict HR for PHC. Therefore, we adopted GM(1,1) model in order to make up for the lack of this research content to a certain extent. This study aims to measure the effect of CHC on the allocation and equity of HR in PHC, at the same time, it provides some real data to support the government’s policies improvement in the next stage.

## 2. Methods

### 2.1 Data resources and region division

The original data on HR in PHC used in our study were obtained from the China Health Statistics Yearbook (2013-2022) (this yearbook collects data from the year prior to the year in which it was published. Its former name includes China Health and Family Planning Statistics Yearbook (2016-2017)) [20]. Data about year-end population size and GDP per capita by province were from the China Statistical Yearbook (2013-2022) [21]. Hong Kong, Macao and Taiwan were not included in this study due to missing data. We divided China into six economic zones, which take into account the geographical location and economic characteristics of each province. Beijing, Tianjin, Hebei, Shanxi and Inner Mongolia are included in the north region; Liaoning, Jilin and Heilongjiang are divided into the northeast region; Shanghai, Jiangsu, Zhejiang, Anhui, Fujian, Jiangxi and Shandong belong to the eastern region; the south-central region includes Henan, Hubei, Hunan, Guangdong, Guangxi and Hainan; the southwest region consists of Chongqing, Sichuan, Guizhou, Yunnan and Tibet; Shaanxi, Gansu, Qinghai, and Ningxia are grouped together as the northwest region. The division of the six economic regions and their GDP per capita in 2021, and some basic information about the healthcare consortium owned by them are shown in the Figure 1.

**Fig. 1.**
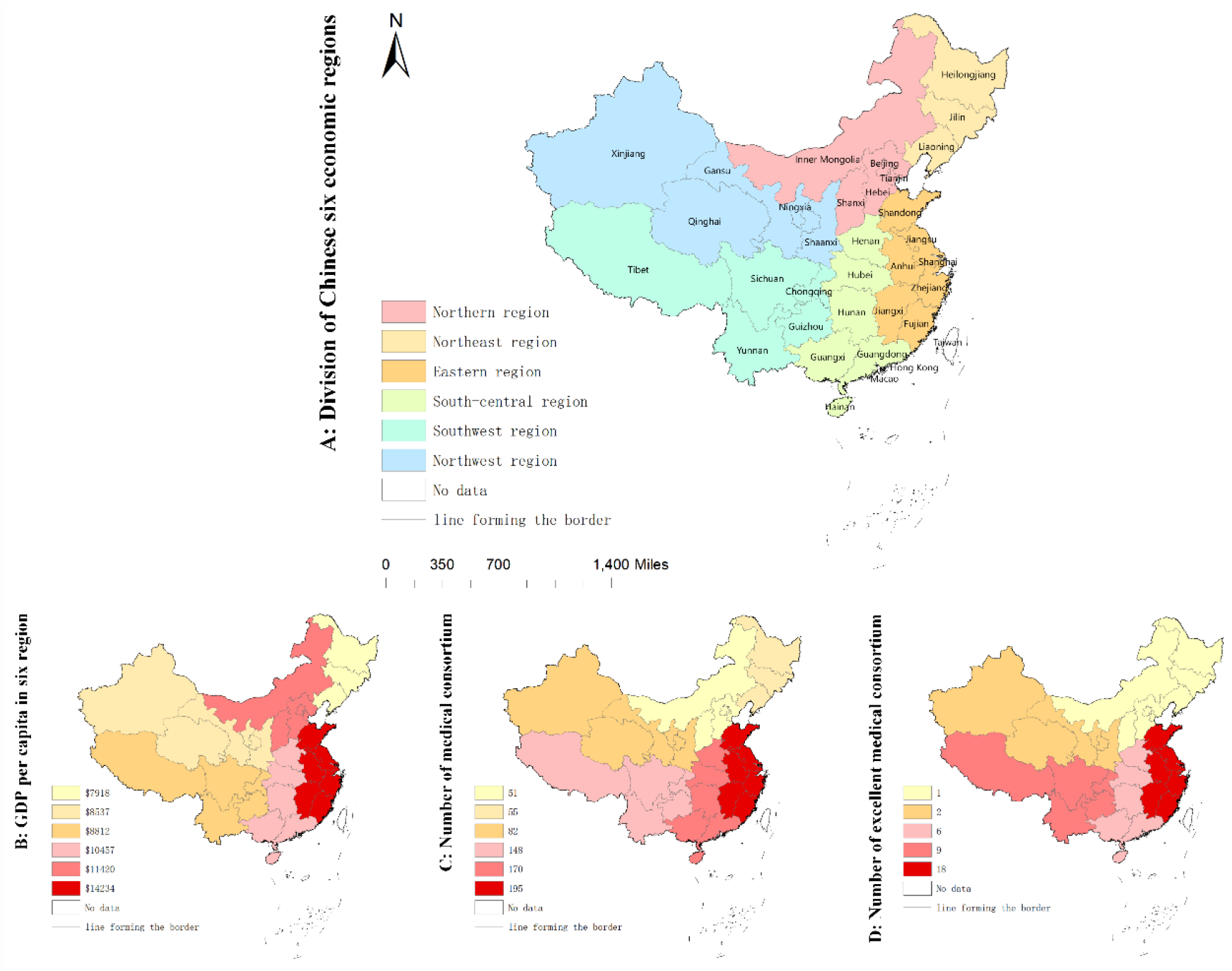
The division of Chinese six economic regions and basic information about them. A shows the six economic regions of China. B shows the GDP per capita of the six regions in 2021. C show the number of pilot sites for medical consortium owned by the six regions in 2019, and D shows the number of medical consortiums owned by the six regions that were rated as excellent in 2021

**Fig. 2.**
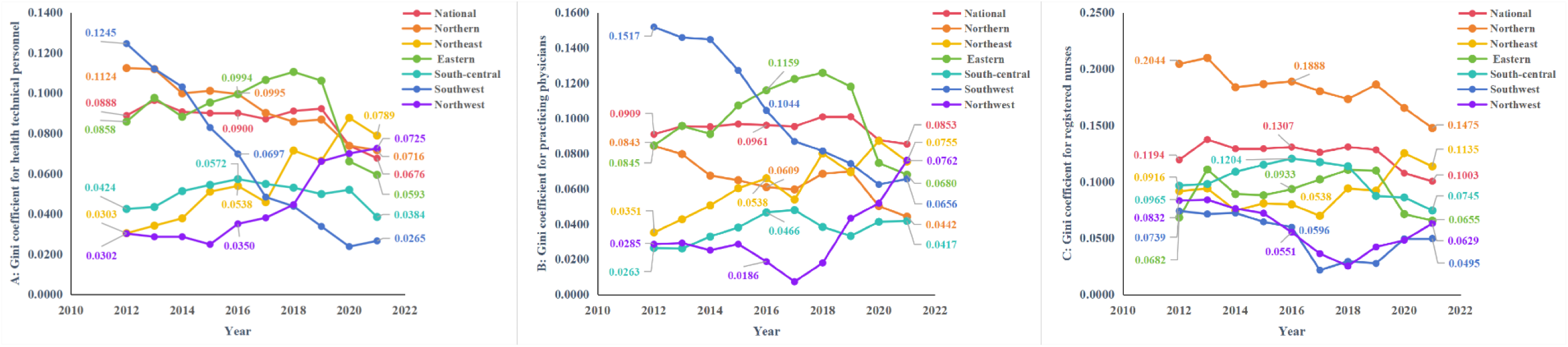
The G of HR for PHC from 2012 to 2021. **A, B**, and **C** show the G of health technical personnel, practicing physicians, registered nurses respectively.

### 2.2 Indicators

Based on the literature review [1,14], three indicators containing health technical personnel, practicing physicians, and registered nurses were selected to assess the allocation of HR in PHC. The reference for the inclusion exclusion criteria of the indicators is the China Health Statistics Yearbook [20].

### 2.3 Gini coefficient (G)

The Gini coefficient, derived from the Lorenz curve, is one of the classic indicators used internationally to measure the degree of inequality in the wealth of the inhabitants of a country or region. G is also often utilized as a measure of equity in the distribution of health resources [14,22]. G is in the range of 0 to 1, G<0.2 denotes absolute fairness; 0.2≤G<0.3 indicates relative balance; 0.3≤G<0.4 means basically reasonable; 0.4≤G represents a relatively high level of inequality; 0.5<G reflects a large inequality gap [23]. Thus, scholars generally consider the grading of the Gini coefficient to be 0.4 is a “cordon sanitaire” for unequal allocation of health resources. G is calculated according to the following formula:

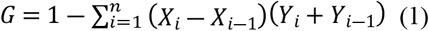

In the above equation, *X*_*i*_ is used to represent the proportion of the population in area of *i* to the overall population size, and *Y*_*i*_ is used to denote the amount of HR for PHC in area of *i* as a proportion of the total amount of these resources.

### 2.4 Theil index (T)

Inequality can also be measured by using the Thiel Index. T has the advantage of breaking down the results into variability between components and variability within components, so that the root causes of inequity can be further located. T has a value between 0 and 1, with smaller values representing greater inequity in different regions. The T is between 0 and 1, with smaller T representing lower levels of inequality in different regions [26]. The formula for T is as follows:

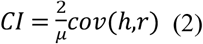

In Eq. (3), *P*_*i*_ is the population of region i as a percentage of the total population, and *Y*_*i*_ is the HR for PHC in region i as a percentage of total resources. T can be further decomposed into *T*_*intra*_ representing intra-regional differences and *T*_*inter*_ indicating inter-regional differences, the details are as follows:

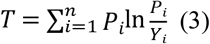

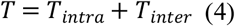

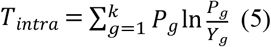

In Eq. (5) and (6), g is used as a proxy for the grouping of the six regions; *P*_*g*_ is utilized to denote the proportion of the regional population to the national population; and the proportion of HR of PHC possessed by the districts to the total national resources is denoted by *Y*_*g*_; m is employed to represent the total number of provinces included within the region; T for each region is denoted by *T*_*g*_ [**Error! Reference source not found**.,14,19]. The values of *T*_*intra*_/*T* and *T*_*inter*_/*T* are referred to percentage contribution to inequality, by which we can tell whether differences in equity originate between or within regions.

### 2.5 Concentration index (CI) and Concentration curve

The concentration index is an evolution of G, through which it is possible to assess the degree of equity in the allocation of health resources under different economic conditions [24]. Concentration curves, on the other hand, are developed from Lorentz curves. There are two ways to calculate CI: Methods of covariance and geometric. The covariance method is computed as follows:

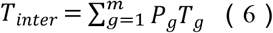

In Eq. (2), *r* is defined as the ordering of GDP per capita, *h* is used as the ordering of the amount of health resources, and *μ* is represented as the mean amount of HR in PHC. In our research, we optioned the geometric method to compute CI, which has the same formula and computational steps as G. However, there are some differences between the concentration curves and Lorentz curves, in that the X and Y axes of the concentration curves correspond to the cumulative percentage of population and the cumulative percentage of HR in PHC after sorting by GDP per capita, respectively. The value of CI ranges from -1 to 1, the closer the absolute value of CI, the better the fairness is. When the CI is negative, the concentration curve appears above the absolute equity line, which implies that more of HR for PHC tend to be allocated to low-income areas. In contrast, it means that more of HR for PHC tend to be allocated to higher-income regions.

### 2.6 Gray predictive model GM(1,1)

GM(1,1) is to establish a first-order continuous differential equation with a time series as the independent variable to predict the future trend of a dependent variable [27,28]. This method has the advantage of long-term forecasting and has been applied to forecast trends in human resources for health care [19]. Our study drew on the computational process of GM(1,1) used by Ruxin Kou [19]. First, the construction of the accumulation matrix and the constant vector are constructed by accumulating the original data sequences. Next, the endogenous control gray level *u* and the development coefficient *a* are calculated. The model equations are then constructed:

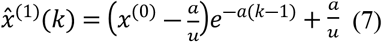

In Eq. (7), *x*^(0)^ represents the original sequence, 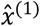 refers to the cumulative sequence that is the original sequence, and *k* is the time series. After the model is constructed, by using a posteriori error ratio C and small error probability P its accuracy will be tested, model accuracy is graded as in Table 1.

**Table 1.** Model accuracy grading scale.

| Predictive accuracy class | posterior variance ratio (C) | posterior probability (P) |
| --- | --- | --- |
| Superior | $\leq 0.35$ | $\geq 0.95$ |
| Qualified | $0.35 < C \leq 0.50$ | $0.80 \leq P < 0.95$ |
| Marginal | $0.50 < C \leq 0.65$ | $0.70 \leq P < 0.80$ |
| Disqualified | $> 0.65$ | $< 0.70$ |

## 3 Results

### 3.1 Temporal changes of the allocation of human resources for primary health care in China from 2012 to 2021

Comparing the three major years, we can visualize that the number of HR for PHC is increasing in both nation and the six economic regions (Table 2). The fastest average rates of growth for health technical personnel and registered nurses are in the Southwest at 6.33% and 11.74% respectively, and practicing physicians is in the Northern at 5.49% (Table 3). Using 2016 as the cut-off point, we compared the average rate of growth in HR for PHC per 1,000 capita in the six regions over the two phases and were surprised to find that the average rate of growth from 2016 to 2021 was significantly higher than that from 2012 to 2016, the average rates of growth of health technical personnel, practicing physicians, and registered nurses in the latter period were 4.01 percentage points, 4.79 percentage points, and 4.18 percentage points higher than in the former period respectively.

**Table 2.**
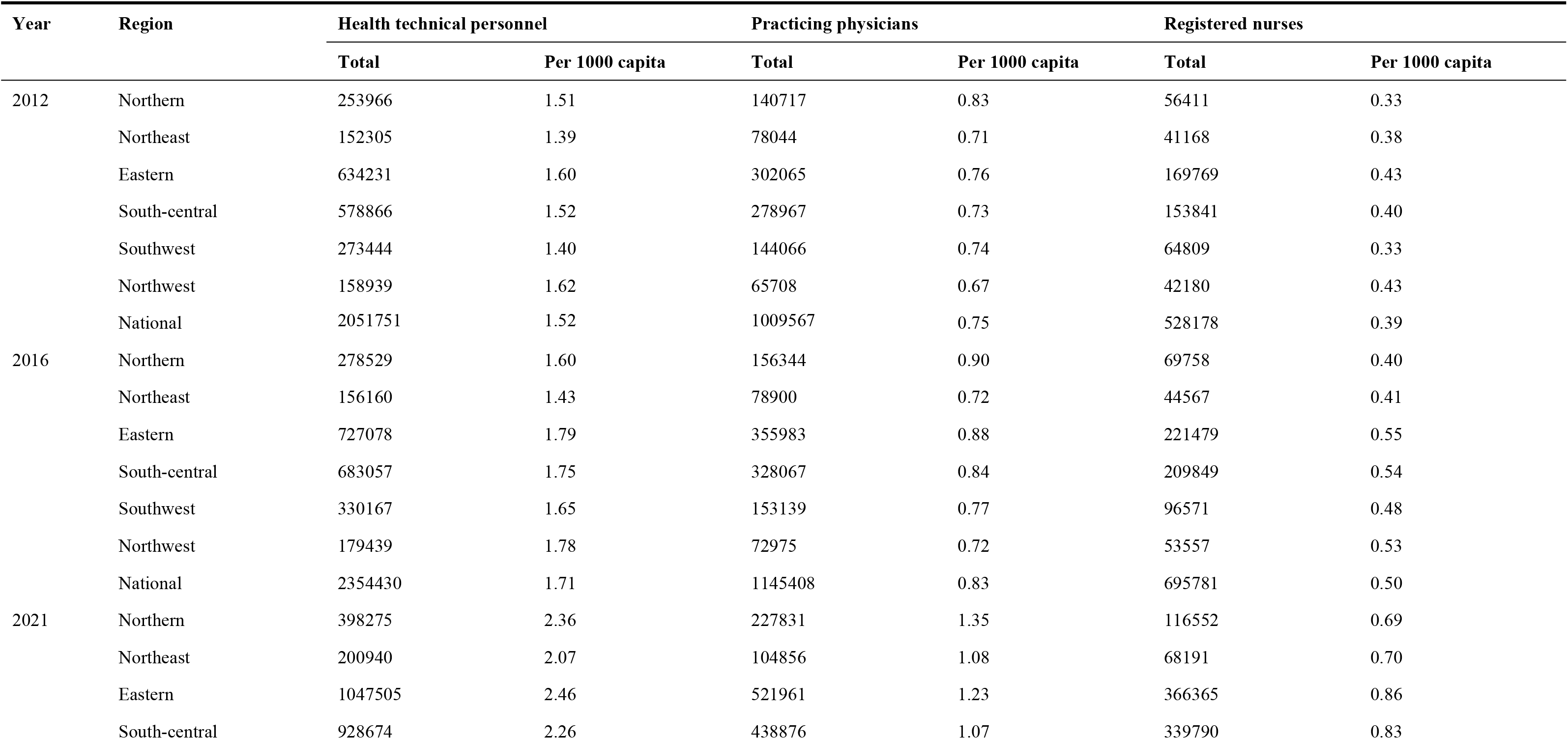
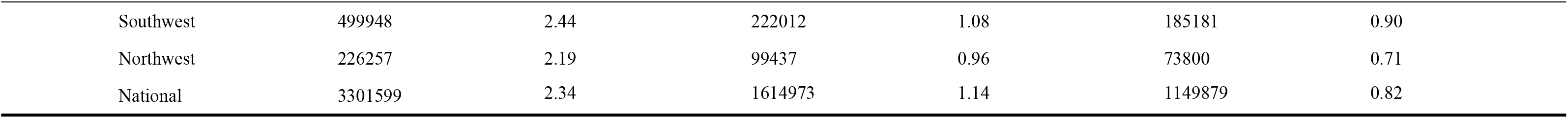
Total and per 1,000 capita of HR of PHC in three major years.

**Table 3.** Average rate of growth of HR for PHC Per 1000 capita by phase.

| region | Health technical personnel |  |  | Practicing physicians |  |  | Registered nurses |  |  |
| --- | --- | --- | --- | --- | --- | --- | --- | --- | --- |
|  | 2012-2016 | 2016-2021 | 2012-2021 | 2012-2016 | 2016-2021 | 2012-2021 | 2012-2016 | 2016-2021 | 2012-2021 |
| Northern | 1.54% | 8.06% | 5.11% | 1.87% | 8.47% | 5.49% | 4.63% | 11.48% | 8.38% |
| Northeast | 0.77% | 7.61% | 4.52% | 0.42% | 8.31% | 4.73% | 2.15% | 11.40% | 7.19% |
| Eastern | 2.89% | 6.60% | 4.93% | 3.60% | 6.97% | 5.46% | 6.27% | 9.58% | 8.10% |
| South-central | 3.55% | 5.24% | 4.48% | 3.46% | 4.90% | 4.25% | 7.37% | 8.98% | 8.26% |
| Southwest | 4.17% | 8.09% | 6.33% | 0.90% | 7.15% | 4.33% | 9.79% | 13.32% | 11.74% |
| Northwest | 2.29% | 4.21% | 3.35% | 1.87% | 5.84% | 4.06% | 5.34% | 6.07% | 5.75% |
| National | 2.99% | 6.47% | 4.91% | 2.57% | 6.55% | 4.76% | 6.41% | 10.40% | 8.61% |

### 3.2 Equity analysis of human resource allocation for Chinese primary health care based on a three-stage approach

#### 3.2.1 Results of the Gini coefficient

The change in G of HR for PHC is shown in Figure 3. At the national level and in the six regions, the G of HR for PHC in the three main years is less than 0.2, and based on this result it can be assumed that there is a good level of equality of allocation for resources. By comparing G from 2012 to 2016 with that from 2016 to 2021 we find that while some regions, such as regions of eastern and south-central, saw their G rose between 2012 and 2016, their inequality levels gradually declining in the years after 2016. There are exceptions, however, the opposite is true in regions of northeast and northwest, where G shown an upward trend in both regions.

**Fig. 3.**
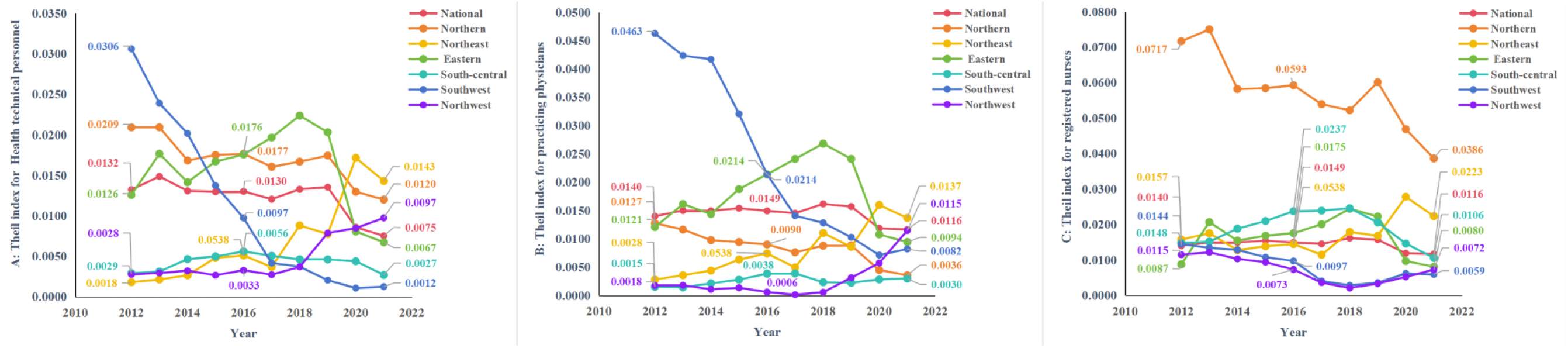
The T of HR for PHC from 2012 to 2021. **A, B**, and **C** show the T of health technical personnel, practicing physicians, registered nurses respectively

#### 3.2.2 Results of the Theil index

The results of analyzing equity through the T are shown in Figure 3, which is essentially the same as the analysis of G. At the same time, Table 4 further shows us the root of the current inequality in allocation of HR for PHC in the country as a whole. It was the intra-regional differences that contributed more to the overall level of inequality in 2021. The percentage contribution of the six regions to intra-region inequality was further calculated, and in Figure 4 it can be observed intuitively that the eastern accounted for the highest percentage of intra-region inequality for health technical personnel and practicing physicians, and the northern accounted for the highest percentage of intraregion inequality for registered nurses. Figure 5 shows the allocation of HR for PHC per 1,000 capita in province in 2021, we can find the reason that the eastern contributed more to the intra-region inequality in configuration of health technical personnel and practicing physicians is that it had both the province with the higher level of allocation and the province with the lowest level of allocation, and the same is true for the northern.

**Table 4.**
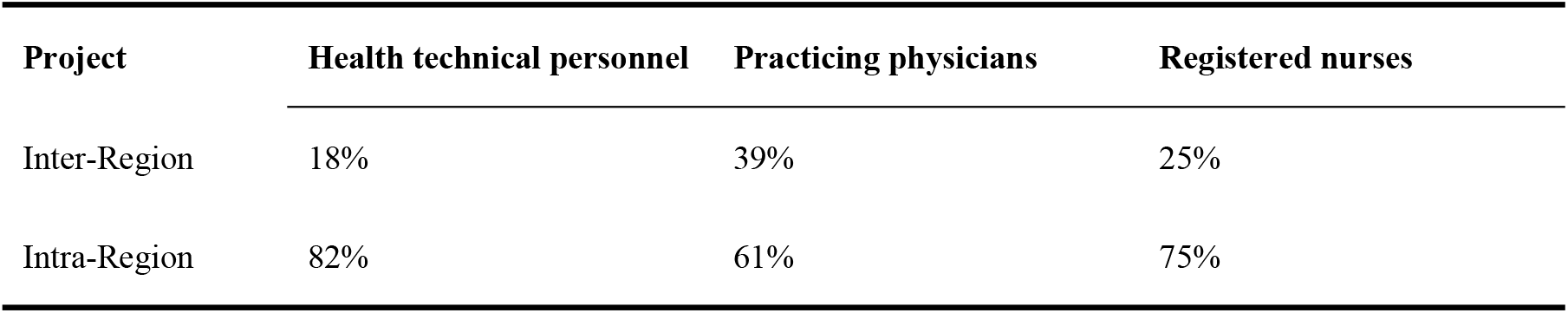
Percentage of inter-regional and intra-regional contributions to T in 2021.

| Project | Health technical personnel | Practicing physicians | Registered nurses |
| --- | --- | --- | --- |
| Inter-Region | 18% | 39% | 25% |
| Intra-Region | 82% | 61% | 75% |

**Fig. 4.**
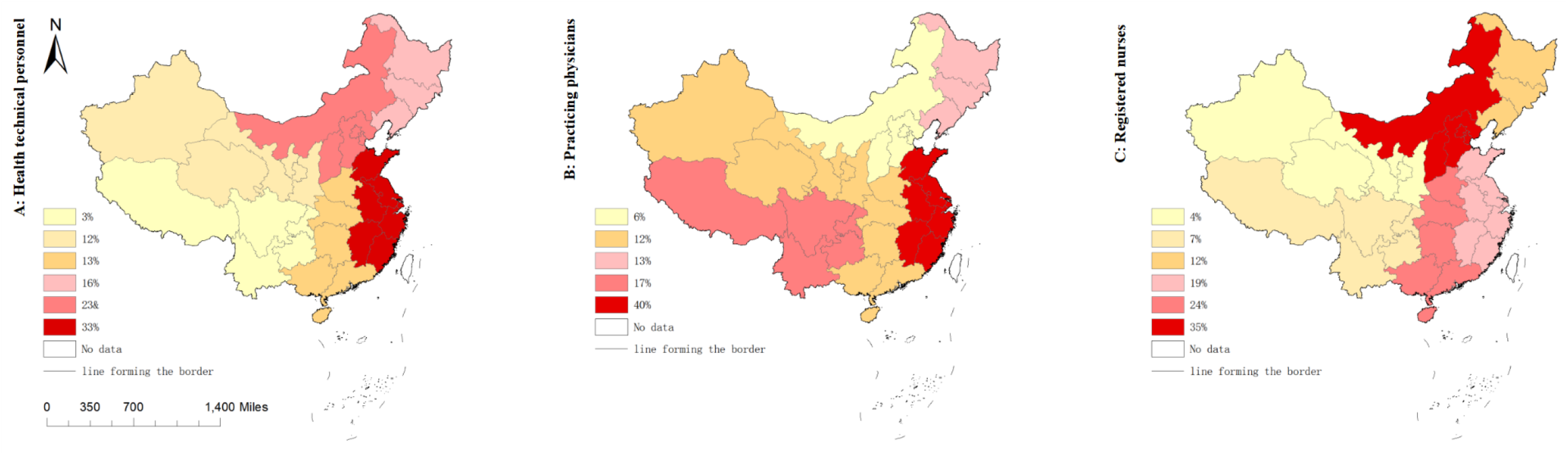
Percentage contribution of the six regions to intra-regional inequality in 2021. **A, B**, and **C** correspond respectively to health technical personnel, practicing physicians, registered nurses

**Fig. 5.**
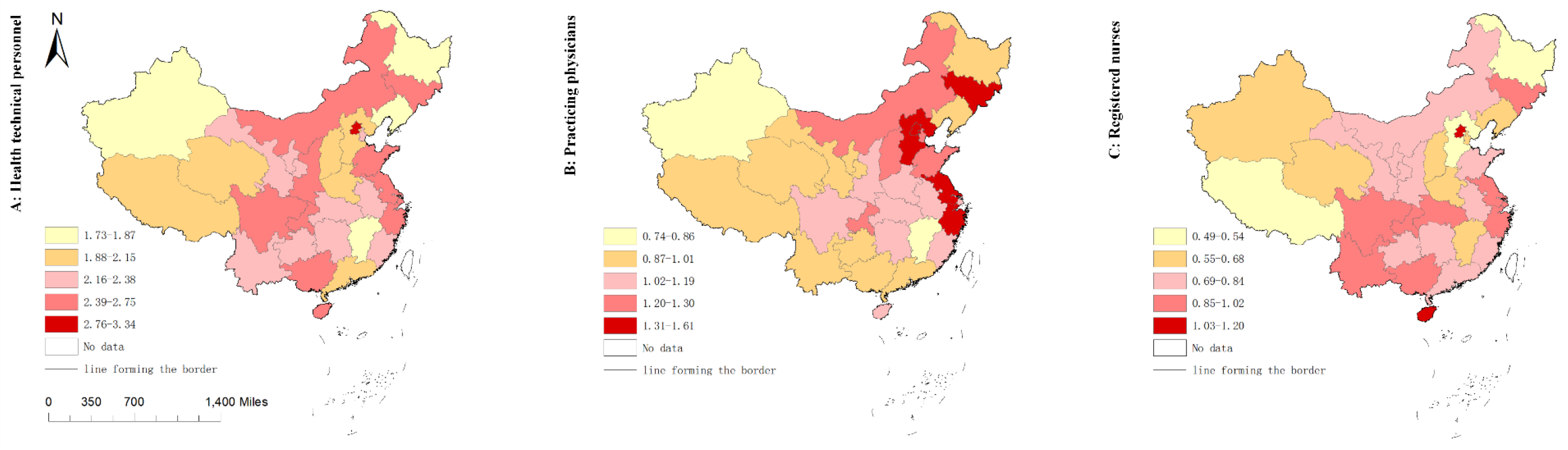
Number of HR for PHC per 1,000 capita in the six regions in 2021. A, B, and C correspond respectively to health technical personnel, practicing physicians, registered nurses

#### 3.2.3 Results of the Concentration index and Concentration curve

The absolute value of the concentration index for all HR for PHC is less than 0.2, indicating a low level of inequality in the allocation of these resources. in concrete terms, in 2012, the overall national CI for health technical personnel, practicing physicians and registered nurses were 0.0407, 0.0458 and 0.0568 respectively. By 2016, their CI increased slightly to 0.0484 for health technical personnel, 0.0594 for practicing physicians, and 0.0657 for registered nurses. But by 2021, their CI have all declined to 0.0302 for health technical personnel, 0.0280 for practicing physicians and 0.0486 for registered nurses respectively. In Figure 6, the changes of inequality in three major years can be seen more visually, with the concentration curves for all resources deviating slightly more outward from the absolute equity curve in 2016 than in 2012, which represents a decrease in their degree of equity between 2012 and 2016. In contrast, the concentration curves for all resources in 2021 were closer inward to the absolute equity curves than they were in 2016, which suggests a change for the better in their equity between 2016 and 2021, and by a comparison of value of CI, equity becomes more improving in 2021 than it was in 2012. What can also be observed from the Figure 6 is that the concentration curve of resources in all years lies below the line of absolute equity, which indicates these resources are more skewed in favor of being allocated to areas with better economic incomes.

**Fig. 6.**
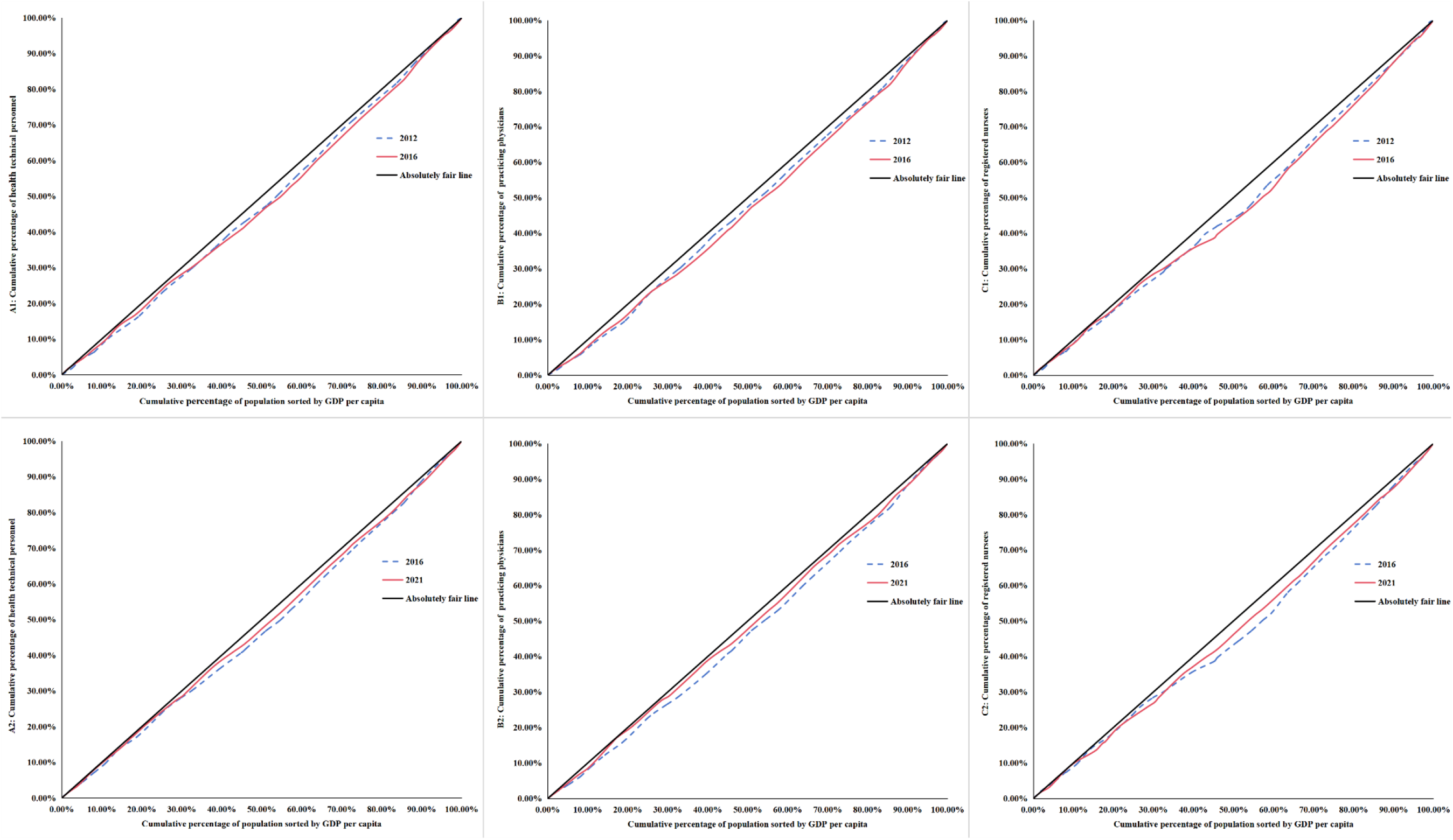
Concentration curve of HR for PHC, 2012 vs. 2016 and 2016 vs. 2021. **A1, B1**, and **C1** show the concentration curve of health technical personnel, practicing physicians, registered nurses respectively, 2012 vs. 2016. **A2, B2**, and **C2** show the concentration curve of health technical personnel, practicing physicians, registered nurses respectively, 2016 and 2021.

#### 3.2.4 Forecasting results of human resources for Chinese primary health care through GM(1,1)model

In this section, we first predicted the total amount of primary health care resources at the national level and in the six districts by constructing a GM(1,1) model. The information underlying the constructed model and the accuracy tests are displayed in Table 5. In the second step, we refered to the data of projected population of Chinese provinces from 2022 to 2030 under the scenario of low fertility accompanied by stable migration in findings of Zhang’s study [30], and combined with the first step, we projected the amount of HR for PHC per 1,000 capita from 2022 to 2030, which is presented in Figure 7. HR for PHC show a growing trend, with health technical personnel, practicing physicians and registered nurses per 1,000 capita reaching 3.88, 1.91 and 1.84 in 2030, but at the same time, we also find that although the total amount of resources in each region is increasing, there are differences in the average rate of growth in each region (Fig. 8).

**Table 5.**
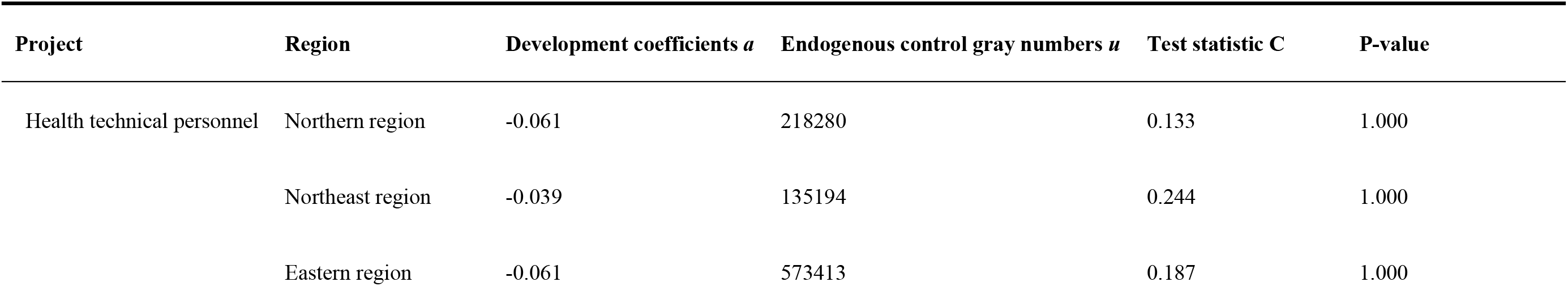
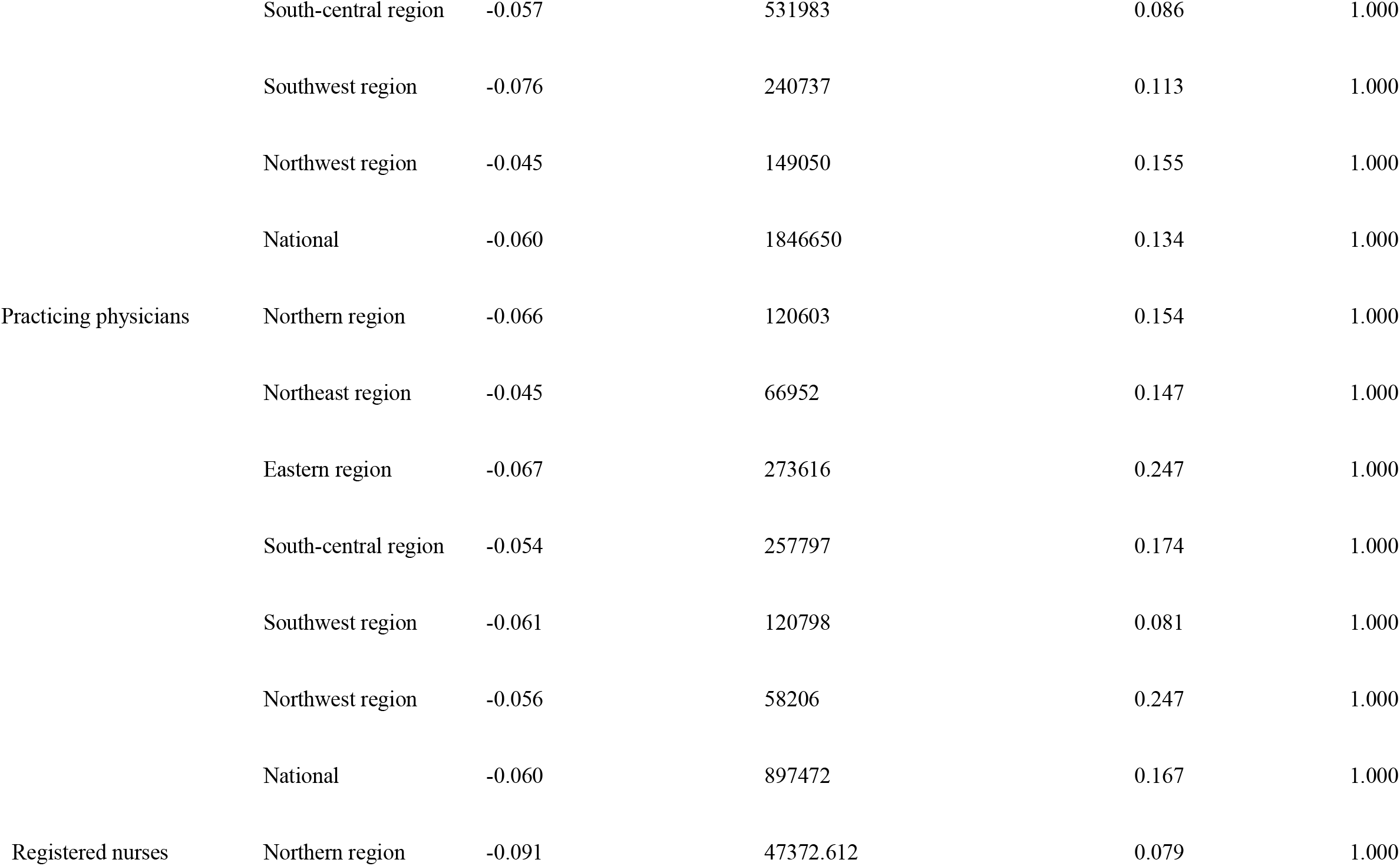
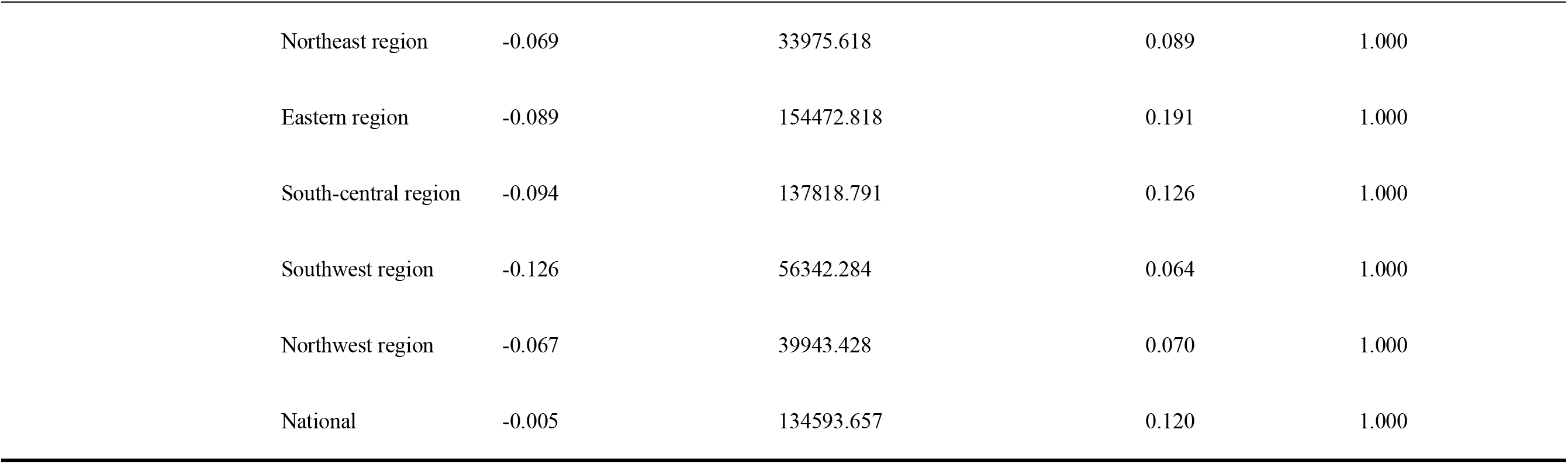
Basic information and value of accuracy test about GM (1,1) model.

**Fig. 7.**
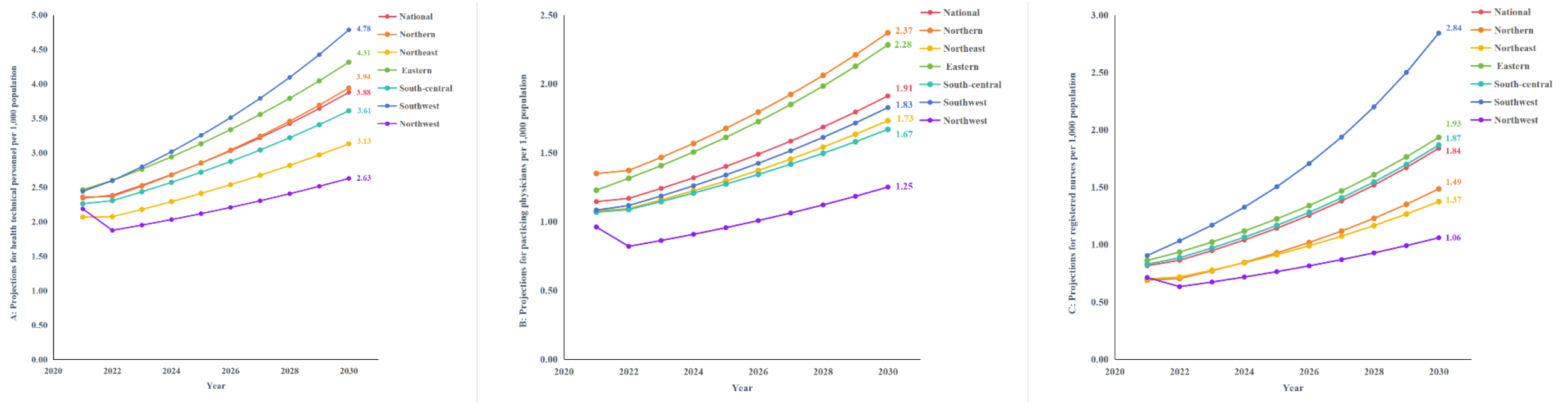
Projections of HR for PHC per 1000 capita from 2022-2023. **A, B**, and **C** show the projections of health technical personnel, practicing physicians, registered nurses respectively from 2022-2023

**Fig. 8.**
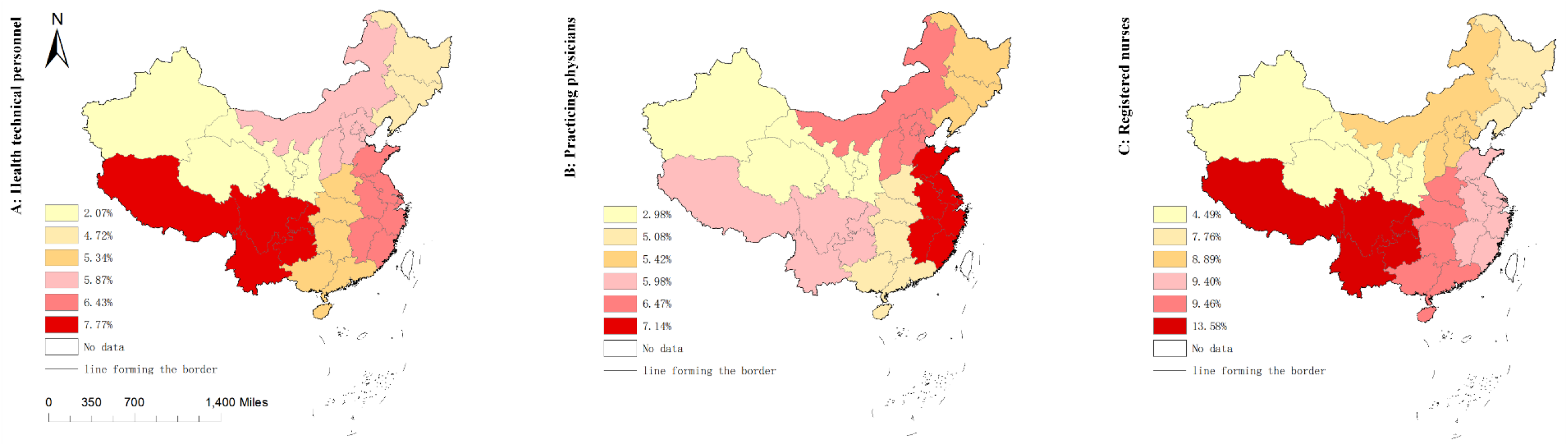
Projected average rate of growth of HR for PHC per 1,000 population from 2021 to 2030

## 4 Discussions

The development of the volume of HR for PHC has been given a stronger boost with the release of CHC in 2016. However, there are differences in the amount of resources allocated and the average rate of growth for resource between regions, and we find that such differences seem to be closely related to the level of economic income and the level of regional implementation of CHC. In the results based on a three-stage fairness analysis of allocation for HR of PHC, the outcomes of the G firstly tell us that these resources are generally equitably allocated by population, and that although inequality increased a bit in some areas between 2012 and 2016, it declined and became more equitable than in the past during the period of 2016 to 2020, similar results can be found in T. The effects of CHC are not always immediate but have a certain lag, this is because the policies have been continuously improved and become clearer in subsequent years, for example, the government released the Guiding Opinions on Promoting the Construction and Development of Healthcare Consortia in 2017 [31], followed by the Work Program for Comprehensive Performance Assessment of Healthcare Consortia (Trial) in 2018 [32], and Measures for the Management of Healthcare Consortia (Trial) in 2020 and so on [33]. These policies have helped localities to continuously clarify the national requirements and measures for the construction of regional healthcare consortia, so their implementation of the policies has become more and more effective, and the influence of CHC on facilitating the equity of allocation of HR in PHC has also been gradually revealed in this process. G also demonstrates the problem that the northeast and northwest regions were not trending toward more equitable allocation of HR in PHC as other regions were, on the contrary, there was an upward trend of G in these two regions after 2016. This may be relevant to the economic income level of the regions as well as the level of policies implemented. By observing at the GDP per capita of the regions of northwest and northeast in 2021, the number of healthcare consortium pilot sites that both regions had in 2019, and the number of healthcare consortia rated as excellent that they had in 2021, both regions were worse than regions with declining inequality trends, such as regions in southwest, south-central, and eastern.

In the analysis of T, in the second part, a further search for the root of inequality is performed. It can be found that the inequality in the total allocation of HR for PHC mainly from intra-region, specifically eastern region is found to contribute more to the inequality in the allocation of health technical personnel and practicing physicians, and northern region is found to contribute more to the inequality in the allocation of registered nurses. By analyzing the allocation of HR for PHC per 1,000 capita in each province of the six regions in 2021, it can be noticed that eastern and northern regions contributed more to the inequality in the distribution of resources is because these two regions include both resource-rich provinces and that with fewer resources, i.e., Jiangxi Province located in the eastern region had fewer health technicians and practicing physicians per 1,000 capita than other provinces in the same district, as does Hebei Province in northern region in terms of registered nurses per 1,000 capita. In 2021, Jiangxi Province and Hebei Province ranked 15th and 27th respectively in terms of GDP per capita in China, while they both ranked last in terms of GDP per capita in their respective regions.

We summarize the phenomena fed back from the analyses of G and T and argue that the allocation of HR for PHC and the equity of its configuration may be affected by both the level of CHC implementation and level of income of the local economy. One potential inference that has been made is that the income level of the economy impacts the total amount of health resources which in turn influences the effectiveness of implementation in policies of CHC which in turn affects the equity of allocation in HR of PHC. According to previous studies, the allocation of health resources is significantly and positively associated with the level of economic income [34], higher-income regions have more financial resources than lower-income regions that can be invested in promoting a more optimized allocation of physical and human resources for health [35], while the human, physical, and financial resources of health collectively form the basis for efficient implementation of the policies in CHC. Based on this, a new question arises: Is it possible that the CHC will aggravate the polarization of HR for PHC in high-income and low-income areas, leading to an increase in the level of inequality?

In order to clarify the above issues, the CI in the third step was performed. In the outcomes of the CI, overall HR in PHC were more inclined to be allocated to higher income areas, but after the implementation of the CHC in 2016, the concentration curve did not expand outward but converged inward towards the absolute equity line, which means that the CHC policy did not exacerbate the degree of differentiation between low-income and high-income areas in terms of inequality in the allocation of human resources in PHC, on the contrary made it has reduced the degree of differentiation. However, we are not sure if the opposite is likely to happen in the future.

In the projections of HR for PHC per 1,000 capita from 2021 to 2030 for the GM(1,1) model with base data from 2012 to 2021, the number of resources will increase further and the average rate of growth comparing 2012 to 2021 for the country as a whole and for most regions from 2021 to 2030 has also accelerated, however, the northwest region’s average rate of growth has slowed and decreases. In 2021, the average difference in the amount of resources between the regions with the best allocation of resources and the regions with the worst allocation of resources is 0.27, but if the current level of development continues, the difference between them will rise to 1.69 by 2030, which seems to indicate that the differences in the distribution of resources between regions are widening. Since the prediction result of the GM (1, 1) model is a natural change without considering external influences [19], in fact, the allocation of HR in PHC will be interfered by complex external factors such as economy, policy, population, public health emergencies [36,37], etc., so our prediction results may not be accurate but it serves as a reminder that it is necessary for the government to pay attention to the CHC’s long-term impact on the inequality of the allocation of HR for PHC.

Four models of healthcare consortiums were proposed in CHC, they are urban hospital groups, rural healthcare associations, cross-regional specialist alliances and remote area tele-collaboration networks, The objective of the first two is to promote the vertical flow of resources within the region, while the latter two emphasize more on improving the allocation of health care resources and the quality of health care services through the horizontal assistance of resource-rich regions with a higher level of health care to resource-poor regions like the northwest regions [13]. A retrospective study noted that the healthcare consortium models enhanced health outputs and health outcomes [38]. In this study, we noticed is the effect of the CHC in improving the allocation and equity of HR in PHC within the respective independent districts, but its impact on reducing disparities between regions is not noticeable, as it does not significantly help areas with a low allocation of resources to become as more resourced as areas with a high allocation.

Based on the above conclusions, it is certain that CHC and its series of sub-policies are of great practical value in achieving improvements in the allocation of human resources for primary health care and their equity in China. It is therefore to be hoped that what the government may consider is to take a long-term view in monitoring the impact of CHC on the allocation of resources to primary health care and the equity of that, and to pay close attention in a nuanced manner to the effects of the implementation of the policy in the regions of different levels of economic income. Because, as the philosophical saying goes, “quantitative change leads to qualitative change”, China’s health care system is a complex system that needs to serve a population of 1.4 billion people, and any minor discrepancies that are not emphasized may in the future be a hidden danger that worsens the allocation of HR for PHC and its equity, which could potentially spill over into the inequality of the population’s health. For the next phase of policy, we suggest that more emphasis should be placed on how to increase the amount of HR allocated to PHC in the backward regions. So how to attract more talents for the backward regions? This may give full play to the advantages of the horizontal assistance model proposed by CHC, whereby the assisted region provides financial resources and policy support, the hospitals that provide assistance export material resources such as medical technology, learning platforms, hospital staffing, etc. and play a well-known role to attract more HR for PHC to resource-poor regions through the establishment of branch hospitals or HR programs.

This study is concerned with analyzing the effects of the implementation of policies of CHC from the perspective of the allocation of the number of PHC human resources by population and its equity, and does not explore the quality of resources and population needs, among other perspectives. At the same time, although we considered the impact of regional economic income and the effectiveness of regional implementation for CHC on the allocation and equity of HR in PHC, we did not take into account other social factors such as the health status of regional populations and educational attainment, thus may have some limitations. The prediction of GM (1, 1) is executed without considering the external factors, so the real result may be somewhat different from the prediction when the external factors change.

## 5. Conclusions

Since the implementation of the CHC, the allocation of HR for PHC in China has improved. Through the three stages of inequality analysis, we conclude that the inequality of HR for PHC in China is low, however, the inequality between regions has not been eliminated. We draw the inference that the level of economic income affects the total amount of health resources, which in turn affects the effectiveness of CHC’s implementation, which in turn affects the allocation of HR for PHC and its equity in China, but of course this interlocking chain of influences needs to be supported by more empirical studies. The GM (1, 1) model shows a growing trend of HR for PHC, but there are differences in the average rate of growth between regions, and the differences in the level of resource allocation may widen in the future. We propose government to take a long-term view to monitor the impact of CHC on the allocation of HR for PHC and its equity, with particular attention to the impact on different economic income regions. At the end we raise an idea to the government about playing the role of the CHC horizontal healthcare consortiums on enhancing HR for PHC in less resourced areas.

## Data Availability

within the manuscript and/or Supporting Information files

## Author Contributions

Conceptualization, S.L.; methodology, S.L.; software, S.L. and Z.L.; validation, S.Z; formal analysis, S.L.; resources, S.Z.; data curation, S.L. and Z.L.; writing-original draft preparation, S.L.; writing-review and editing, S.L.; visualization, S.L. and Z.L.; supervision, S.Z.; project administration, S.Z. All authors have read and agreed to the published version of the manuscript.

## Informed Consent Statement

Not applicable.

## Availability of data and materials

The datasets used and/or analyzed during the current study are available from the corresponding author on reasonable request.

## Institutional Review Board Statement

The data are publicly available data. Therefore, it was considered an exempt category.

## Ethical approval

Not required.

